# Attitudes Towards Coronavirus (COVID-19) Vaccine and Sources of Information Across Diverse Ethnic Groups in the UK: a Qualitative Study

**DOI:** 10.1101/2022.02.04.22270456

**Authors:** Eirwen Sides, Leah Ffion Jones, Atiya Kamal, Amy Thomas, Rowshonara B Syeda, Awatif Kaissi, Donna M Lecky, Mahendra G Patel, Laura B Nellums, Jane Greenway, Ines Campos-Matos, Rashmi Shukla, Colin Stewart Brown, Manish Pareek, Loretta Sollars, Emma Pawson, Cliodna AM McNulty

## Abstract

**Objectives:** To explore attitudes and intentions towards COVID-19 vaccination, and influences and sources of information about COVID-19 across diverse ethnic groups (EGs) in the UK.

**Design:** Remote qualitative interviews and focus groups (FGs) conducted June-October 2020 before UK COVID-19 vaccine approval. Data were transcribed and analysed through inductive thematic analysis.

**Setting:** General public in the community across England and Wales.

**Participants:** 100 participants from 19 self-identified EGs with spoken English or Punjabi.

**Results:** Mistrust and doubt were common themes across all EGs including white British and minority EGs, but more pronounced amongst Bangladeshi, Pakistani, Black ethnicities and Travellers. Many participants shared concerns about perceived lack of information about COVID-19 vaccine safety, efficacy and potential unknown adverse effects. Across EGs participants stated occupations with public contact, older adults and vulnerable groups should be prioritised for vaccination. Perceived risk, social influences, occupation, age, co-morbidities and engagement with healthcare influenced participants’ intentions to accept vaccination once available; all Jewish FG participants intended to accept, while all Traveller FG participants indicated they probably would not.

Facilitators to COVID-19 vaccine uptake across all EGs included: desire to return to normality and protect health and wellbeing; perceived higher risk of infection; evidence of vaccine safety and efficacy; vaccine availability and accessibility.

COVID-19 information sources were influenced by social factors, culture and religion and included: friends, family; media and news outlets; and research literature. Participants across most different EGs were concerned about misinformation or had negative attitudes towards the media.

**Conclusions:** During vaccination programme roll-out, including boosters, commissioners and vaccine providers should provide accurate information, authentic community outreach, and use appropriate channels to disseminate information and counter misinformation. Adopting a context-specific approach to vaccine resources, interventions and policies and empowering communities has potential to increase trust in the programme.

**Article summary: strengths and limitations:**

- This is amongst the largest qualitative studies on attitudes to the COVID-19 pandemic in the UK general public across ethnic groups (EGs), ages and religions, adding insights from a broader range of participants.
- Qualitative methodology enabled discussion of participants’ responses around COVID-19 vaccination, probing to collect rich data to inform recommendations across EGs.
- Most data collection was undertaken in English, possibly excluding sectors of the population who may access COVID-19 information through different sources due to language.
- Data collection was June-October 2020 before COVID-19 vaccines were licensed. Attitudes are highly responsive to current information around a COVID-19 vaccine, as well as the state of the pandemic and perceived risk. Data were collected prior to much of the intervention work, putting the attitudes and intentions expressed in this study in a context of minimal community engagement and support. This provides a baseline snapshot of attitudes, providing the option to explore and assess the impact of such interventions.
- Socioeconomic data and index of multiple deprivation were not collected, limiting the ability to determine a possible accumulative effect of factors such as socioeconomic status, ethnicity and age.

## Introduction

The coronavirus (COVID-19) pandemic has had a striking impact on global health with five million reported deaths worldwide by December 2021[1].The UK has seen more COVID-19 cases per capita than many other countries with over 13 million cumulative cases up to January 2022, over 172,000 deaths and much associated morbidity including ‘long COVID-19’[2]. Increased COVID-19 morbidity and mortality have been associated with increasing age, gender, comorbidities, deprivation, occupations with greater face to face contact, and certain minority ethnic groups (EGs)[3-5].

Vaccination programmes are one of the key strategies used to limit the societal impact of infections[6] so vaccine acceptability and uptake is crucial to COVID-19 control[7]. Public vaccine safety concerns and doubts have contributed to reductions in uptake of non-COVID-19 vaccines which has led in increase in these infections[8, 9].

Modelling suggests 10,400 deaths and much long-term morbidity had been avoided by March 2021 through the English COVID-19 vaccination programme introduced in December 2020 [10]. Positive COVID-19 vaccine attitudes reportedly increased from 78% in December 2020 to 96% in May to June 2021[11, 12].

Evidence indicates there were differences in COVID-19 vaccine uptake based on demographic and socioeconomic factors. Black or Black British adults and those living in the most deprived areas were more likely to report COVID-19 vaccine hesitancy[11-14]. However, there is a lack of in-depth qualitative literature exploring attitudes towards the COVID-19 vaccine across a broad cross-section of the UK population with balanced representation of ethnicities, ages, genders and religions. This qualitative study aims to explore the general public’s acceptability and uptake of a COVID-19 vaccine prior to its roll out and attitudes towards sources of COVID-19 information, with representation across EGs.

## Methods

This paper forms part of a wider qualitative study that explored the attitudes, behaviours and needs of diverse EGs in Wales and England during the COVID-19 pandemic.

### Focus group and interview topic guide development

The interview guide (Supplementary 1) was informed by Public Health England’s (PHE) 2020 review of disparities in risks and outcomes for COVID-19[4] and the Theoretical Domains Framework (TDF)[15]. Eight of the fourteen TDF domains were mapped to the following topics explored in the interviews and FGs: knowledge, beliefs about consequences of COVID-19 infection and vaccination; optimism that a vaccine would help solve the pandemic (optimism); feelings about being offered a COVID-19 vaccine (emotion, memory, attention and decision-making); reasons for or against accepting a COVID-19 vaccine (intentions, reinforcement); influences on decision-making (memory, attention and decision making); groups to be prioritised (professional role and identity); and where to receive the vaccine if willing (environmental context and resources).

### Recruitment

Individuals were recruited between June and October 2020 with the aim of selecting a diverse cross-section of the UK population across regions, including minority EGs, religions, and occupations.Ethnic minority varies by country and context and is defined as “a group of people who differ in race or colour or national, religious, or cultural origin from the majority population of the country in which they live”[16]. Around 80.5% of the population of Wales and England belongs to the White British EG[17]. Minority EGs include: Asian EGs 7.5% (including 2.5% Indian, 2.0% Pakistani, 0.8% Bangladeshi and 0.7% Chinese), Black EGs 3.3% (including 1.8% Black African and 1.1% Black Caribbean), Mixed/Multiple EGs (2.2%), White 4.4% other, White Irish (0.9%) White Traveller (0.1%) and other EGs (1.0%)[17].

Participants were recruited via a range of methods including: adverts posted on Facebook groups related to COVID-19 support for minority EGs; Twitter; PHE’s People’s Panel; charities who aim to empower and advocate for minority ethnic communities and improve their access to services); and snowball sampling, a method whereby existing study participants refer further participants[18]. The advert (Supplementary 2) requested individuals from diverse ethnic backgrounds, with a reasonable level of spoken English or Punjabi(South Asian language) to participate in 60-minute remote focus groups (FGs) about their experiences during the COVID-19 pandemic. Participants were offered a £25 contribution to thank them for their time.

### Data Collection

Data were collected between 15 June–1 October 2020, prior to first Medicines and Healthcare products Regulatory Agency (MHRA) COVID-19 vaccine approval in the United Kingdom and start of the rollout of the vaccination programme in December 2020[19]. FGs were conducted in English and3 interviews in Punjabi, all via Skype (some participants dialled in over the telephone and video function was optional). FGs were led by one researcher (LJ), supported by a research assistant (ES or RBS). Field notes made during the FGs by research assistants (RBS and ES) were reflected upon with the facilitator (LJ). Interviews in Punjabi were led by one researcher (AK). Only participants and researchers were present at data collection. During the data collection, the topic guide was used flexibly. Participants had the option to join FGs with or without video and discussions lasted approximately 60 minutes, were recorded, transcribed verbatim by an external agency, checked for accuracy by the research team and translated from Punjabi to English where necessary. Findings were discussed weekly by researchers and four times with the study steering group. Data collection stopped once it was agreed by the steering group that a range of ethnic and religious groups had been recruited and data saturation had been reached.

### Data analysis

Transcripts were analysed inductively using thematic analysis in QSR NVivo[20] by three researchers (LJ, AK and ES). A fourth researcher (AT), double coded 12 of 27 transcripts to check consistency. A coding consensus was reached between the four researchers through meetings and discussions. Themes were identified from the data in two internal meetings, one halfway through analysis and another at the end of analysis. Themes were presented, discussed three times with the steering group and finalised in a steering group workshop. Representative quotes were chosen to demonstrate the themes.

### Research Team

The research team, consisted of two researchers (LJ and AK), supported by four research assistants (ES, RBS, AT and AWK), led by two senior researchers DL and CAMM. Researchers and steering group included Arab, British Bangladeshi, British Pakistani, White British and White Irish EGs. The research team was advised by the steering group (including public and healthcare professional (HCP) representatives, MGP, LN, JG, ICM, RBS, CB, MP, LS, EP. Researchers and steering group members were experienced in qualitative research, behavioural science, intervention development, public health, health psychology, and minority ethnic health.

### Patient and Public Involvement

A member of the public was involved in the study steering group from the conception of the study. They also inputted into the design and methodology, as well as data collection tools and recruitment strategy.

### Ethics

The study was internally reviewed by the PHE Research Ethics and Governance Group (REGG) (Reference: NR0215). All participants involved in the study provided informed consent, including the use of anonymised transcript quotes in reporting and publications.

## Results

Initially data from different EGs were analysed separately, however as there were few differences between ethnicities, data are reported for all participants together. Any differences are highlighted.

### Participant Characteristics

141 participants were approached, 100 of whom participated in the study. A total of 24 FGs were conducted in English and 3 interviews in Punjabi, all via Skype.Participants represented a mix of self-reported ethnicities, ages, religions, genders and UK regions, as shown in Table 1, Supplementary Material 3.

**Table 1.**
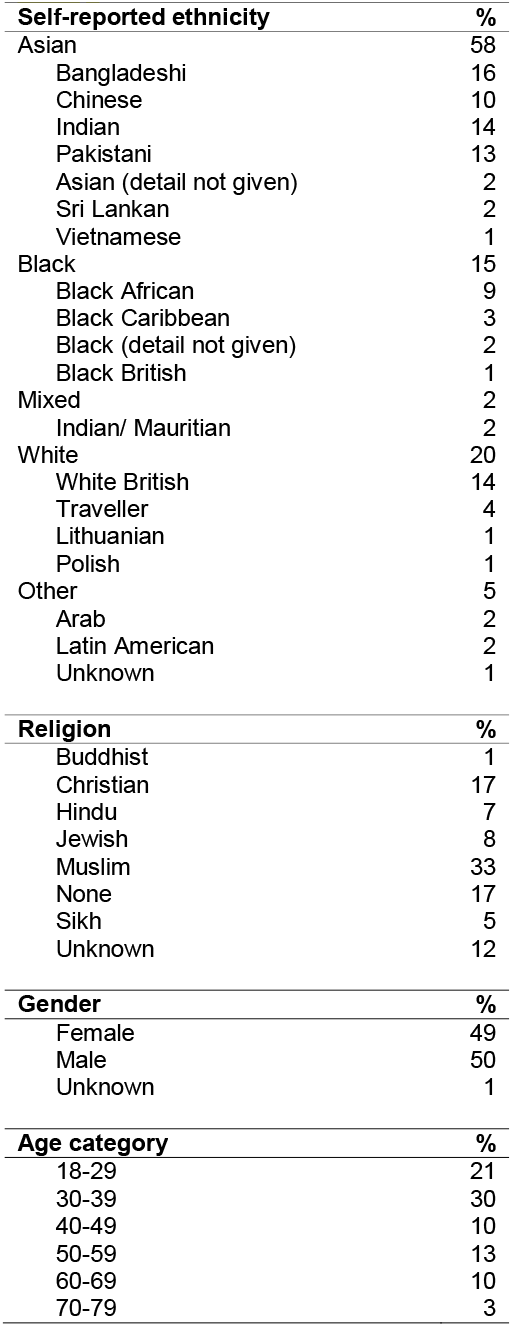
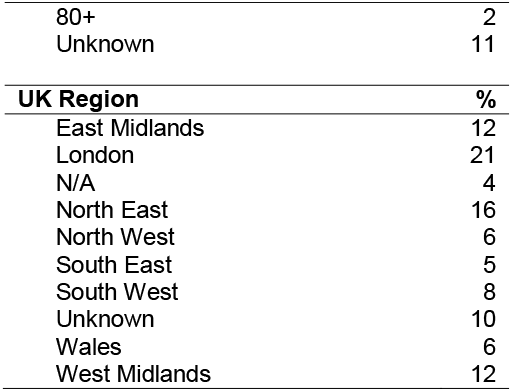
Participant characteristics (n=100)

### COVID-19 vaccination uptake

Three main themes were identified relating to vaccine uptake included: (1) attitudes and beliefs towards COVID-19 and a COVID-19 vaccine; (2) facilitators; and (3) barriers. Subthemes and relationship to the TDF[15] can be viewed in Table 2.

**Table 2.**
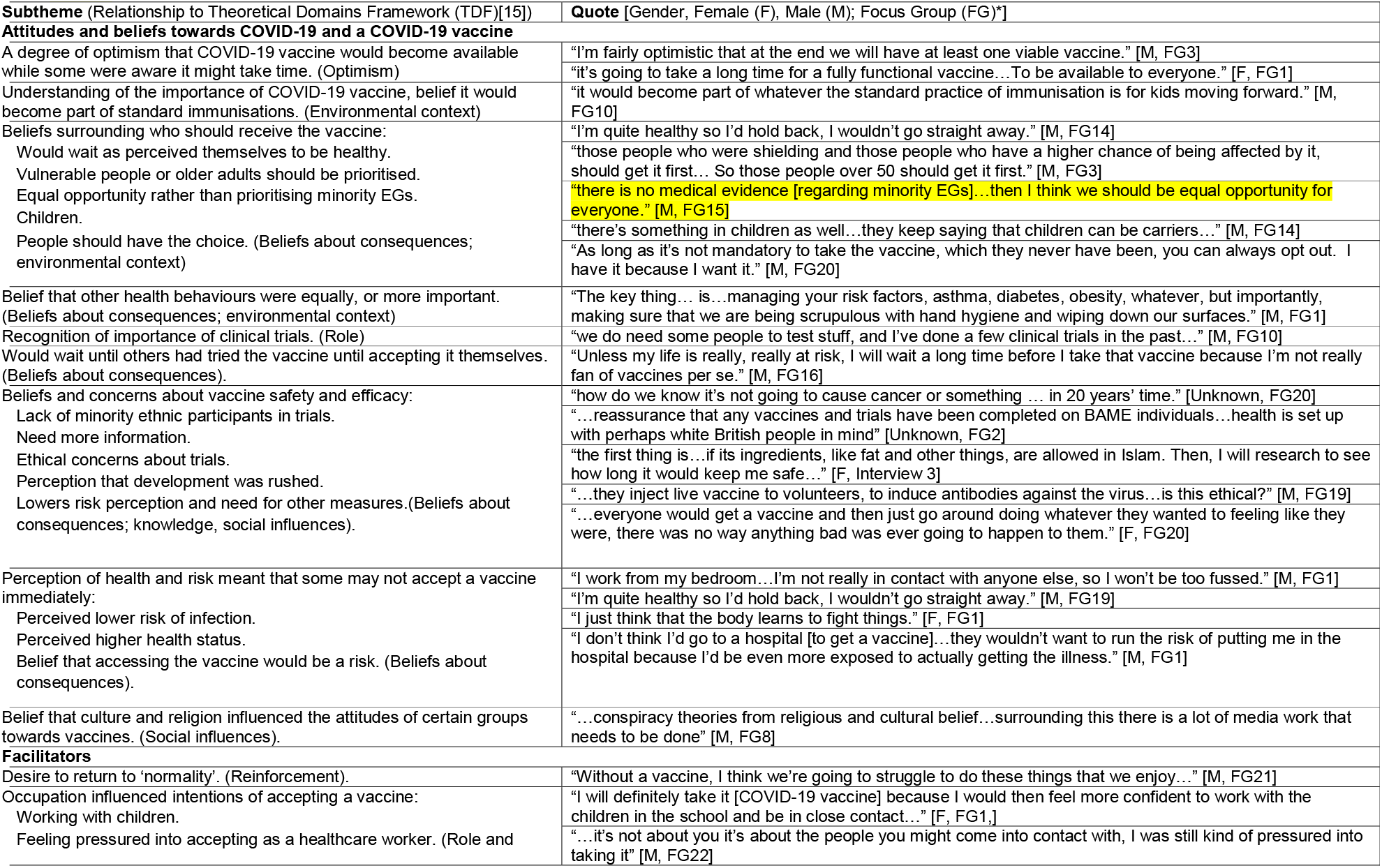
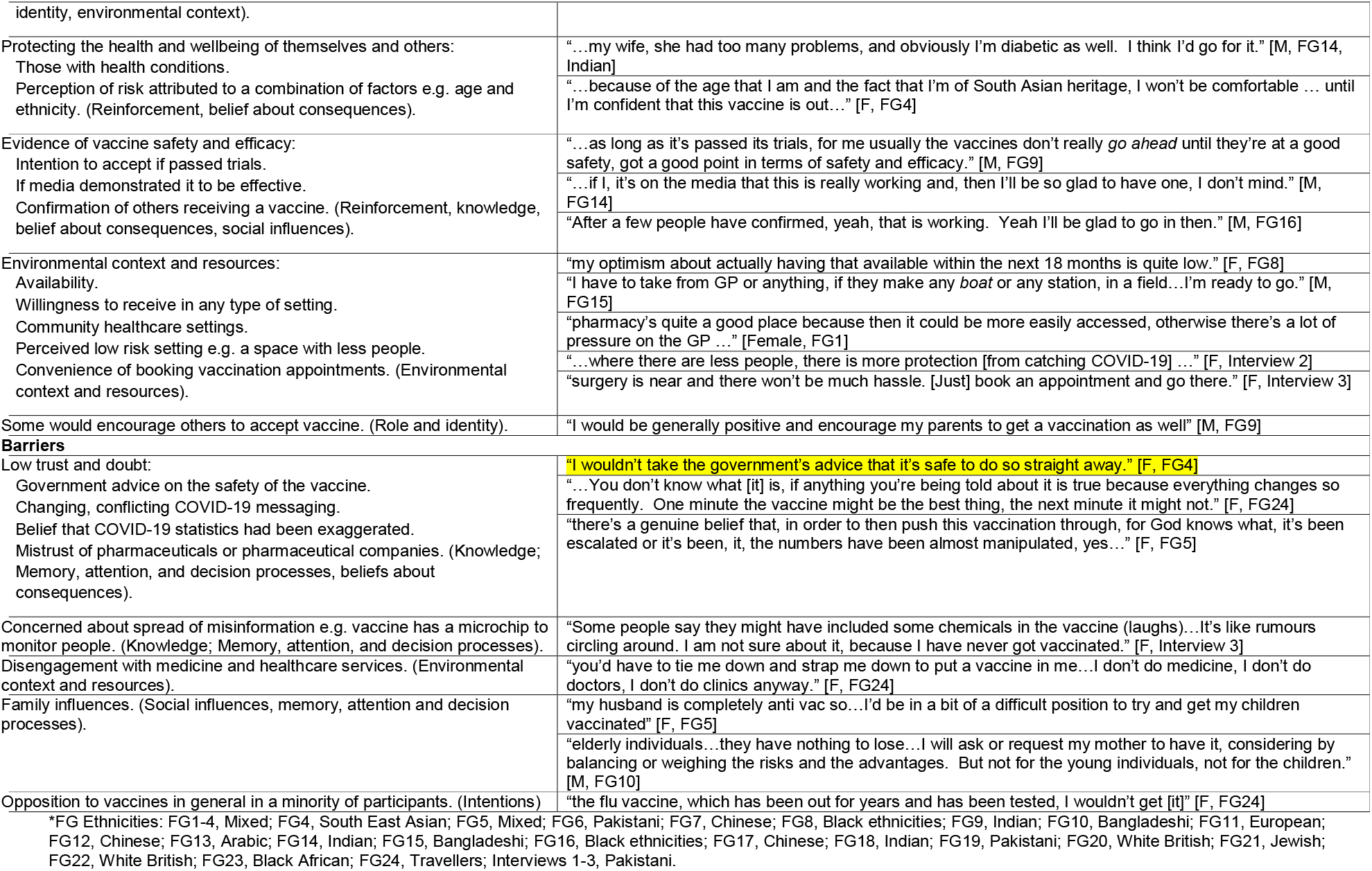
COVID-19 vaccination uptake: attitudes, beliefs, facilitators and barriers.

Within all EGs participants stated mixed intentions about the likely future uptake of COVID-19 vaccine, ranging from full intention to vaccinate to no intention at all to vaccinate. All participants in the Jewish FG intended to accept a vaccine while all Traveller FG participants reported that they probably would not.

### Attitudes and beliefs towards COVID-19 and a COVID-19 vaccine

Across EGs most people (even some of the vaccine hesitant) held similar beliefs on priority groups for vaccination, including occupations with public contact, older adults and vulnerable groups. Another view shared across EGs was that they would not want to be the first to have the vaccine due to concerns about vaccine safety, efficacy and unknown side effects. Beliefs and concerns about COVID-19 vaccine safety and efficacy were raised. One participant wanted reassurance that a vaccine had been trialled amongst ethnic minorities.

There were some differences of opinion on whether children should be prioritised; some thought children should be prioritised as they were carriers of the virus, while others raised concerns about possible unknown side effects on developing immune systems. More detailed findings by ethnic group are reported in Table 3.

**Table 3.**
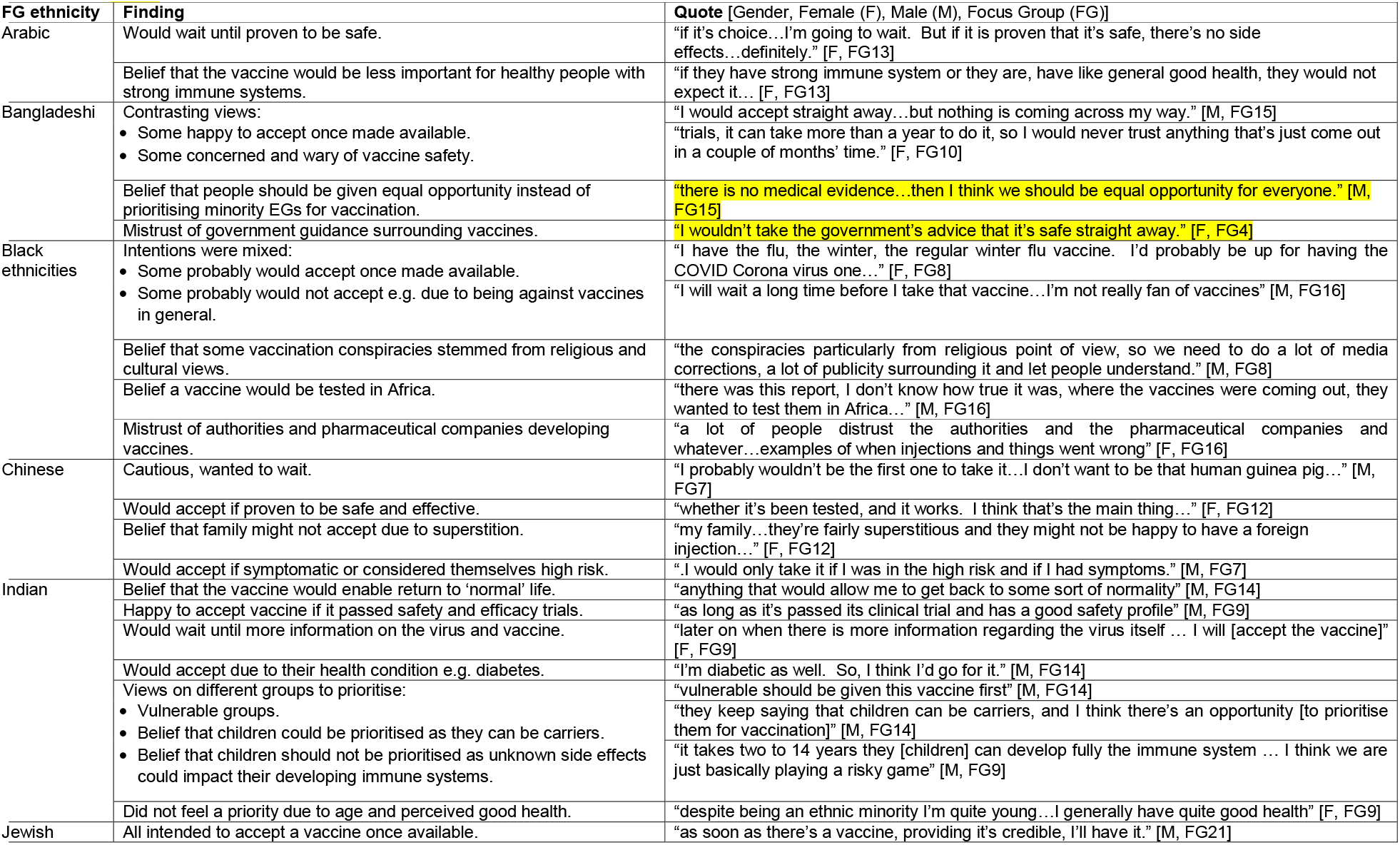
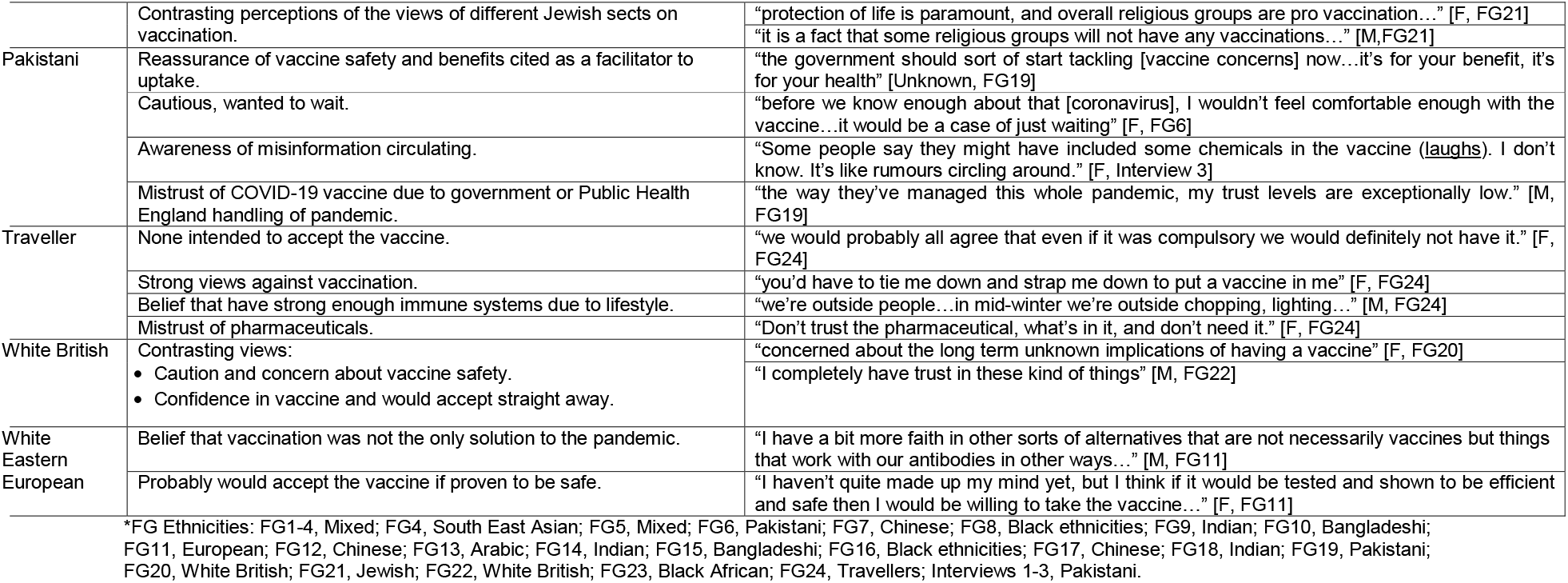
COVID-19 vaccination attitudes and intentions by ethnic group.

There were also differences within EGs, with factors such as frontline occupation and perceived health status influencing their intention to accept a vaccine once made available. Having a health condition led to higher risk perception while positive health status caused lower risk perception, thus influencing intention to accept a vaccine. There was a belief amongst some that alternative methods of prevention such as practicing good hygiene, maintaining a good diet and exercise, were equally, if not more, important to vaccination. Some believed that there were other ways to build immunity rather than vaccination or had a belief that vaccination was unnecessary due to their perception that they were healthy.

Some participants were optimistic that a working vaccine would become available while others were aware it might take time. Some believed a vaccine to be important and recognised its role in herd immunity.

### Barriers to vaccination

Mistrust and doubt were common themes across EGs and many shared concerns about perceived lack of information about COVID-19 vaccine safety, efficacy and potential unknown adverse effects. Mistrust in government advice and recommendations were identified as the greatest potential barriers to vaccine acceptability. This was due to perceptions of the government’s handling of the pandemic, perceived unclear messaging and frequently changing guidance at various stages of the pandemic which resulted in confusion. Disengagement with pharmaceuticals, medicine and healthcare services was a barrier to vaccine uptake which was mainly due to mistrust. Some participants raised concerns about their friends and family being susceptible to misinformation, for example that the vaccine included chemicals or microchips. A few participants had negative views around vaccination imposed by their relatives. A minority of participants stated that they would definitely not accept the vaccine, which was primarily due to being opposed to vaccines in general.

### Facilitators for vaccination

There was general agreement across EGs on places to receive the vaccine, including community healthcare settings and settings perceived as low risk, e.g. a space with less people. Many participants stated that they would accept the vaccine either to enable return to normal life, to continue working, or to protect themselves and others due to existing health conditions. However, several of these participants stated that they would wait until others in the population received the vaccine first in order to observe potential side effects.

### Attitudes and intentions by ethnic group

Caution and wariness to accept a COVID-19 vaccine, including concerns about vaccine safety was reported amongst all EGs including White and non-White groups. Although themes of mistrust and doubt arose across most EGs, they were more pronounced amongst the following: Bangladeshi (mistrust of government guidance surrounding vaccines); Pakistani (mistrust of COVID-19 vaccine due to government and Public Health England handling of pandemic); Black ethnicities and Travellers (mistrust of authorities, pharmaceuticals or pharmaceutical companies developing vaccines).

Arabic and Traveller FG participants suggested that vaccines might not be necessary for those with a strong enough immune system. Some participants in all EGs indicated they probably would accept a vaccine once it became available. There were no other notable differences between EGs.

### Sources of COVID-19 information

Themes around COVID-19 information included: (1) sourcing information from friends, family and social media, media and news outlets and the research literature (2) concerns about misinformation; and (3) cultural and religious influences (Table 4).

**Table 4.**
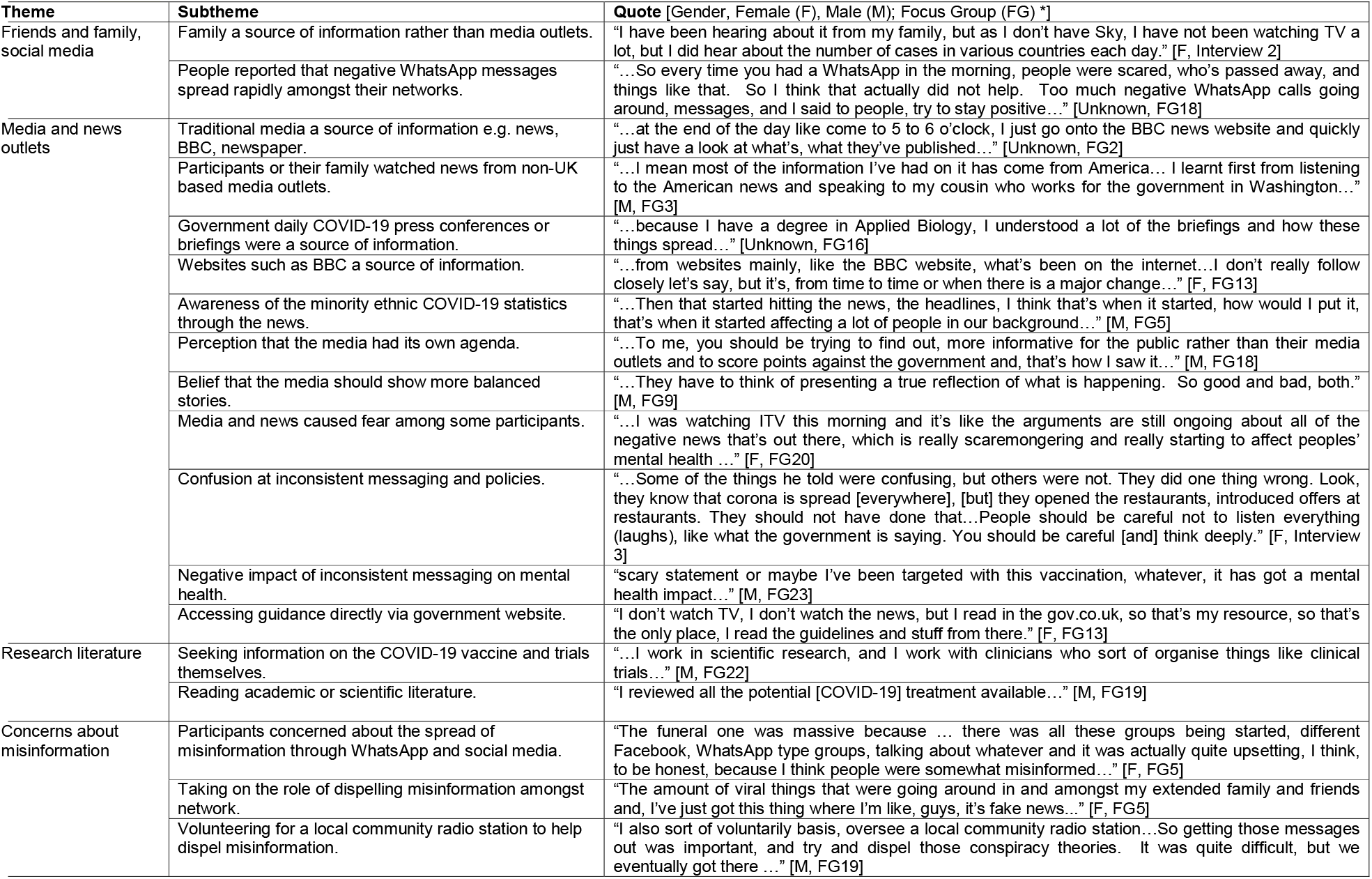
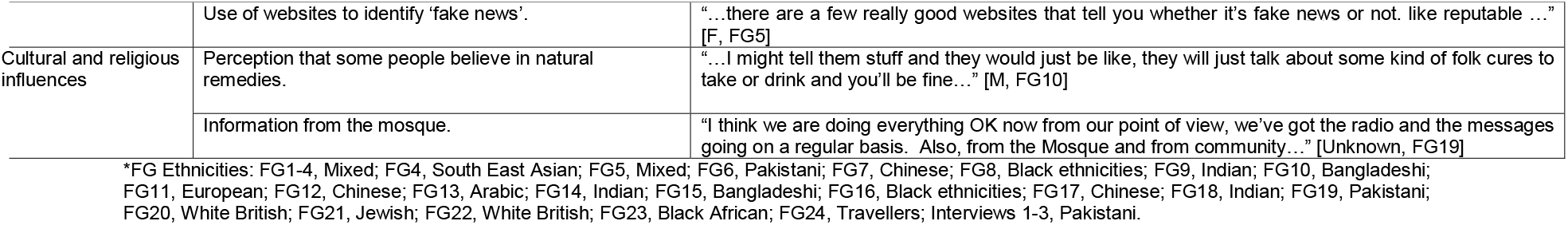
Sources of COVID-19 information

Many participants across all EGs reported comparing stories with friends and family, often via WhatsApp and other social media channels. A number of participants stated that they received information from traditional UK media channels such as British Broadcasting Company (BBC) news and their websites. Some participants reported watching the BBC government daily COVID-19 briefings while others used websites to obtain information and reported that they were aware of the minority ethnic COVID-19 statistics through the news. A small number reported that some relatives obtained information through non-UK based news outlets (e.g. American and Asian), which may have promoted different information, behaviours, and attitudes.

In most focus groups or interviews across a range of EGs, one or more participants had negative attitudes towards the media. Such attitudes included a belief that media: had its own agenda; should present more balanced stories; caused confusion; gave inconsistent messaging, and participants reported wariness or uncertainty surrounding the credibility of the information. It was reported that media coverage had negative implications on the mental health and wellbeing of some participants, sometimes causing fear and distress.

A minority reported directly using government guidance. Some had public facing roles and they therefore followed the guidance from their workplace. A minority reported researching the topic themselves through research literature.

Concerns around misinformation were mentioned across all EGs. Some raised their concerns surrounding the spread of misinformation amongst their WhatsApp and social media networks, for example that the vaccine contained a microchip to monitor people. As a result, some reported taking on the role of dispelling misinformation circulating among friends and family, particularly for older family and community members.

Some cultural and religious sources of information were identified. A participant suggested that their parents believed in traditional remedies (Bangladeshi FG) while another reported obtaining information from the mosque (South East Asian FG).

## Discussion

### Statement of principal findings

This study adds findings about COVID-19 vaccinations, some of which differ from attitudes towards other vaccinations. Mistrust and doubt surrounding COVID-19 vaccination were common themes that arose across white British and minority EGs but they were more pronounced in Bangladeshi, Pakistani, Black ethnicities and Travellers. Across EGs, many were cautious and shared concerns about COVID-19 vaccine safety and efficacy. There were differences within EGs, with factors such as occupation and perceived health status influencing their intention to accept a vaccine once made available. Attitudes and intentions sometimes differed between EGs, for example all participants in the Jewish FG reported that they probably would accept a vaccine while all participants in the Traveller FG probably would not accept one. Although many received their information from trusted sources such as mainstream television, many also reported negative attitudes towards the government, media and news outlets. Table 5 provides an overview of practical intervention and policy recommendations based on the findings of this study.

**Table 5.**
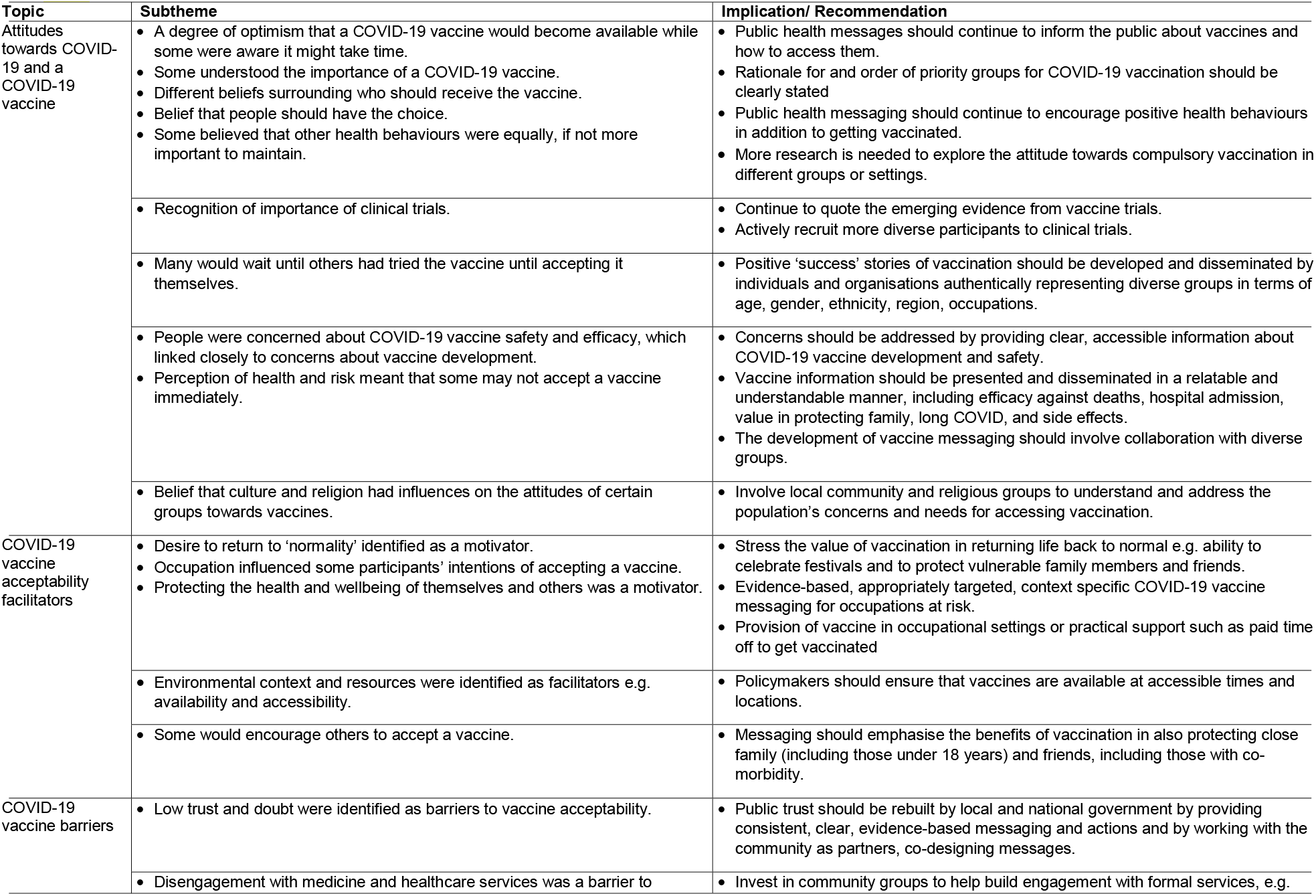
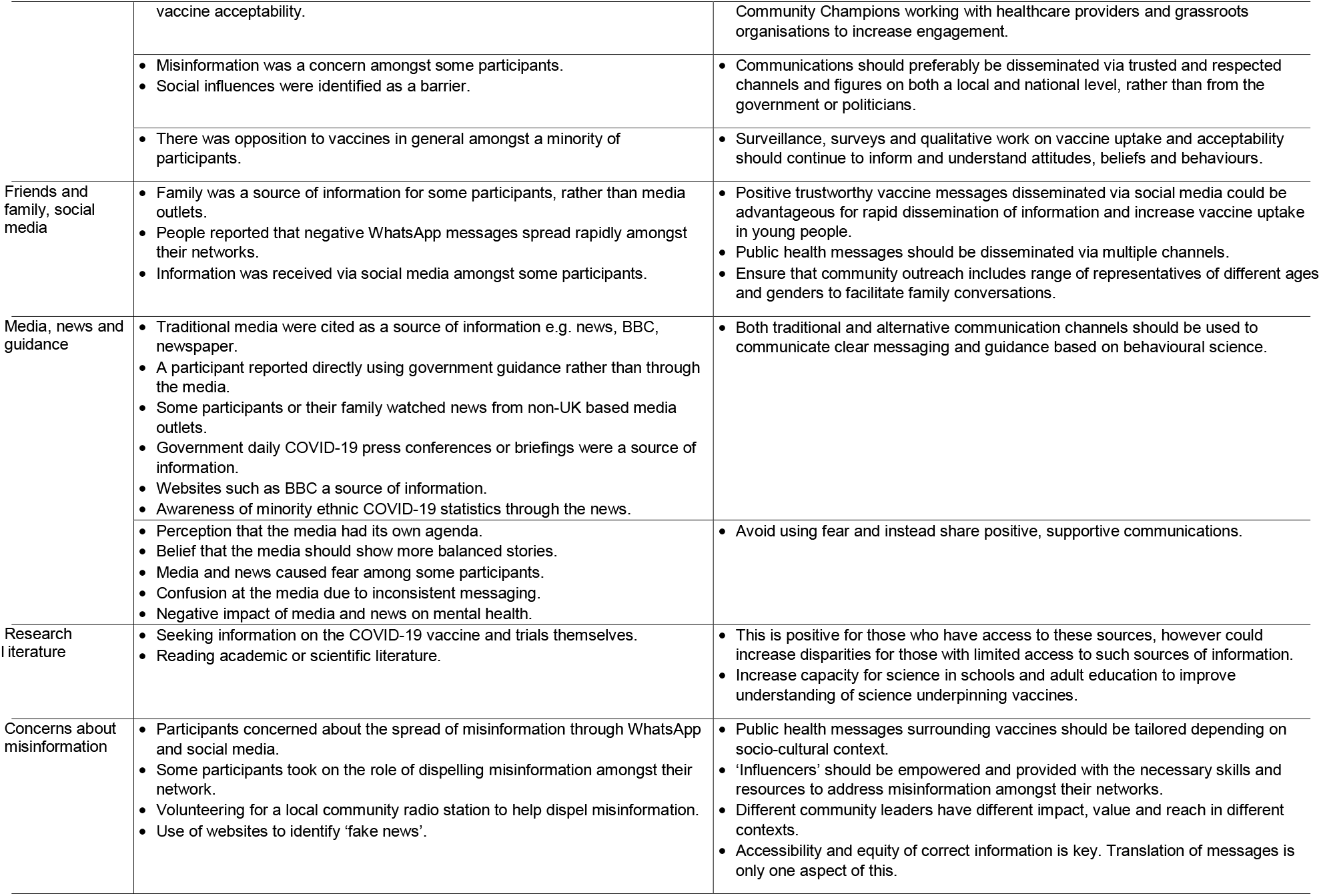
COVID-19 vaccination attitudes and COVID-19 sources of information: implications and recommendations for clinicians and policymakers

### Comparison with existing literature

Low trust in government advice and recommendations due to its perceived handling of the pandemic and changing COVID-19 messaging was identified as a potential barrier to vaccine acceptability and uptake in our study and others [21] [22]. The link between mistrust in a COVID-19 vaccine and mistrust in government was found to be more pronounced among some minority ethnic participants in a small qualitative study amongst UK HCPs, particularly Black African, Black Caribbean and other Black groups[23]. Such differences were observed to an extent in our study, where mistrust was reported in not only Black ethnicities (authorities and pharmaceutical companies developing vaccines), but Bangladeshi (mistrust of government guidance surrounding vaccines) and Pakistani (mistrust of COVID-19 vaccine due to government and Public Health England handling of pandemic) groups too. It is important to recognise that mistrust can stem from wider inequalities beyond COVID-19[23]. In our study, similar barriers and facilitators towards the COVID-19 vaccination were often reported across EGs. However, surveillance data demonstrates that COVID-19 vaccination rates in the UK and Israel were lowest amongst certain minority EGs[24] [25]. For UK healthcare workers between December 2020 and February 2021, studies found that some minority EGs were more likely to be COVID-19 vaccine hesitant in comparison to White British groups[13], and that COVID-19 vaccine uptake was lower amongst some ethnic minority groups compared to White[26]. In a US youth survey, Black participants were less likely and Asian participants more likely to accept a COVID-19 vaccine compared to White participants[27]. A UK survey found that certain demographic characteristics (age, sex, ethnicity, religion, qualifications, employment status, key worker, extremely clinically vulnerable, extremely clinical vulnerable household member) explained only 4% of the variance in vaccination intention, beliefs about the value of vaccines explained 35% of the variance, and positive COVID-19 vaccination beliefs and attitudes explained 28%[28]. This suggests, in line with our findings, that being from a minority EG alone is unlikely to account for vaccine uptake differences and implies that attitudes and intentions vary depending on an interaction between multiple factors such as location, time and socioeconomic, cultural and political context. Locally appropriate outreach settings are needed with flexible appointments to overcome vaccine access issues. Migrants with precarious immigration status suggested walk-in centres in trusted locations such as foodbanks, community centres and charities would facilitate vaccine access and uptake[29]. Additionally, allowing vaccination without documentation or general practice registration should be considered and publicised to facilitate equitable access, for example for the estimated 600,000 undocumented migrants living in the UK,[30] the homeless and other vulnerable populations.

In most focus groups or interviews across a range of EGs, one or more participants had negative attitudes towards the media and its impact on them. In our study, participants raised concerns about the impact of social media misinformation and stress the importance of disseminating clear, consistent vaccination messages covering safety, effectiveness, as well as empowering the public to address misinformation in their networks and being active on social media to combat misinformation (Table 5). Negative attitudes towards the media, government, medicine and healthcare could be overcome by messaging and vaccine delivery from trusted figures [31]. Other studies have found a strong correlation between a trusted healthcare professional or physician’s recommendation of a vaccine and higher uptake[32] [33]. However, this may not be adequate for those who are disengaged with medicine and healthcare. Many participants expressed concerns about receiving a COVID-19 vaccine or wanted more information, particularly around safety and efficacy. Large UK questionnaire surveys support this, demonstrating a significant positive association between confidence in the importance, safety and effectiveness of a COVID-19 vaccine, and vaccine acceptance [21, 34]. A small qualitative UK parental study and larger survey completed in May 2020 found that COVID-19 vaccine safety and efficacy concerns were the greatest barrier to definite vaccine acceptance, which in the larger parallel survey was 56% [21]. There was a belief that COVID-19 vaccine development had been rushed amongst most participants in our study and other small qualitative studies amongst UK HCPs[23]. Some of our participants stated that they would wait until “it is deemed safe and effective,” or others in the population received the vaccine first before accepting it themselves. This was echoed in UK qualitative studies exploring COVID-19 vaccination in pregnant women[35] and recent migrants [29], a Canadian qualitative study in a diverse sample of the population [36] and a US quantitative study of attitudes towards COVID-19 vaccination in 14 to 24 year olds[27].

Previous research indicates that people deem older vaccines safer than newer ones [37, 38]. Main reasons for hesitancy were concerns about unforeseen future side effects of vaccines, and general mistrust in the benefits and safety of vaccines[34], and a few thought that they should rely on their own strong immune systems. A large UK parental survey found that lower income, or ethnic minority participants were at least twice as likely to reject COVID vaccination[34] and although we found no apparent differences by minority ethnicity, our sample size of 100 and its qualitative methodology were not designed to determine this. In a qualitative study on experiences of participation in COVID-19 vaccine trials, minority ethnic participants highlighted the importance of diverse representativeness in trials[39], which was also mentioned by one of our participants. Similar barriers and facilitators to ours were found in a systematic review exploring parents’ acceptance of childhood vaccinations in general [40].

We found that participants’ perception of risk of COVID-19 infection and severe illness to themselves, and their family, through occupation, age or comorbidity, and protection through vaccination were strong facilitators for COVID-19 vaccination acceptance. This has been found in several other studies of the general population, healthcare workers, immunocompromised and parents[7, 21] [32]. In contrast, Travellers in our study and others perceived themselves at lower risk of infection through their lower contact with other population groups.[32] Easy access will be important to facilitate vaccination uptake for those with risk due to occupation or comorbidity[24]. Our study and others certainly indicated that many would prefer a local, low risk community healthcare setting with convenience of booking appointments[29, 41, 42]. However, a key difference to note is that at the time of data collection, access to vaccines was not a tangible issue as they had not yet been approved. The importance of differentiating between vaccine hesitancy, which has less variation in different EGs[43], and under-vaccination related to environmental context and access have been raised by others[29, 44]. In Israel, low uptake in certain groups has been increased by well-tailored outreach efforts[25, 45].

Many participants across EGs reported comparing stories with friends and family, often via channels such as WhatsApp and other social media. Concerns about misinformation were raised across all groups. People with more of a reliance on social media and social networks for COVID-19 information are more likely to trust it [46], and be exposed to misinformation [47]; these tend to be those of younger age, lower education levels, and lower income. [48] There is evidence of social media outlets circulating COVID-19 misinformation.[46, 49-51] [52]. COVID-19 vaccine messaging must be appropriate for both the ‘influencers’ and the ‘influenced’ to facilitate the dissemination of trusted information amongst networks.

## Strengths and limitations of the study

This is amongst the largest qualitative studies on attitudes to vaccination in the UK general public and, importantly in contrast to others, incorporates most UK minority EGs, the COVID-19 pandemic and perceived risk.[7, 21, 24, 25, 28, 34, 43, 48, 53-55] Although intention does not necessarily directly translate to actual vaccine uptake in the future it is a good predictor and surveys demonstrate a steady increase in vaccine acceptance since 2020[53]. Most data collection was undertaken in English, possibly excluding sectors of the population who may access COVID-19 information through different sources due to language. Similar themes were identified from the English FGs and Punjabi interviews, with exception of some religious views, indicating consistency of results. Much of the data collection and analysis was conducted by White British researchers which could have impacted interpretation of findings, however FGs and interviews were held remotely which may have reduced this and also acquiescence bias. The first five FGs included a range of ethnicities while others mainly comprised participants of the same ethnicity; the latter allowed greater reflexivity between participants. Both focus group type yielded similar data. Data collection was June-October 2020 before COVID-19 vaccines were licensed. Attitudes to vaccine are not static and are highly responsive to current information around a COVID-19 vaccine, as well as the state of the pandemic and perceived risk. Data were also collected prior to much of the intervention work, putting the attitudes and intentions expressed in this study in a context of minimal community engagement and support. This is a strength as it provides a baseline snapshot of attitudes, providing the option to explore and assess the impact of such interventions. To avoid exclusion of typically underrepresented groups, recruitment involved approaching charities that aim to empower and advocate for minority ethnic communities and improve their access to services. The data may be subject to selection bias, as those with a greater interest in COVID-19 may have volunteered and we did not reach every minority EG in the UK. Socioeconomic data and index of multiple deprivation were not collected, limiting the ability to determine a possible accumulative effect of factors such as socioeconomic status, ethnicity and age. Attitudes and intentions by EG were presented for the FGs and interviews that included participants of the same or similar ethnicities i.e. FGs 4, 6-24 and Interviews 1-3, as transcription did not allow for differentiation between ethnicities of each participant (Table 3). Table 1 and Supplementary 3 demonstrate FG characteristics.

### Implications for clinicians and policymakers

Interventions and policies must be appropriate and effective for diverse populations where vaccine acceptability and uptake are low, to reduce inequalities and increase vaccine equity. This study’s findings have local and national implications for clinicians and policymakers, as presented in Table 5, which fall under three overarching areas: providing information that addresses specific concerns of communities; authentic community outreach; and using the right channels to disseminate credible information and counteract misinformation.

### Unanswered questions, future research and implications

Since this work was completed the results and recommendations have been presented to government bodies. Faith-based and EG communities are now more actively involved in local and more tailored COVID-19 communications in the UK[56]. There are efforts to locate vaccination clinics in more accepted local assets, such as places of worship, including mosques and churches[56]. Local COVID-19 vaccine community champions and influencers in minority groups are being identified and encouraged[56-58]. Further detailed guidance from the UK Race Disparities Unit encourages targeted local action and engagement with support from community champions and other local leadership[59, 60].

Nonetheless, more high-quality research and evaluation is needed to demonstrate the effects of different interventions on COVID-19 vaccine uptake[5]. Future locally led outreach should engage marginalised groups and explore the attitudes and behaviours where there is low vaccine uptake to mitigate barriers[14]. Future research must gain further understanding of similarities and differences within groups to adopt a context-specific approach to vaccine resources, interventions and policies, and proactively involve diverse patient and public groups. Such interventions should provide access, equity, and knowledge, and empower and engage local communities. Surveillance should continue to monitor vaccine uptake, with both quantitative and qualitative studies to explore any ongoing disparities in uptake and whether they continue to be related to concerns in vaccine safety or low perception of COVID-19 risk.

## Data Availability

Data available upon reasonable request.

## Acknowledgements

We would like to extend our thanks to all public representatives, healthcare professionals, researchers and expert advisors who contributed to this study. Thank you to all participants for providing their time and sharing their views and experiences for FGs and interviews.

## Contributors

- ES: assisted with data collection; had substantial contributions to the analysis and interpretation of the qualitative data; drafted all versions of the manuscript and critically revised it; gave final approval of the version to be published; and has agreed to be accountable for all aspects of the work.
- LJ: project managed from study start to close; led the analysis and interpretation of the qualitative data; had substantial contributions to the design of the study (led on development of protocol, gained ethics approval, drafted interview questions, recruited participants); led the collection of data; had substantial contributions to the analysis and interpretation of the qualitative data; critically commented on versions of the manuscript; gave final approval of the version to be published; and has agreed to be accountable for all aspects of the work.
- AK: had substantial contributions to the design of the study (commented on interview questions, recruited participants); collected data; had substantial contributions to the analysis and interpretation of the qualitative data; critically commented on versions of the manuscript; gave final approval of the version to be published; and has agreed to be accountable for all aspects of the work.
- AT: had substantial contributions to the analysis and interpretation of the qualitative data; critically commented on versions of the manuscript; gave final approval of the version to be published; and has agreed to be accountable for all aspects of the work.
- RBS: had substantial contributions to the design of the study (development of protocol, drafted interview questions); collected data; critically commented on versions of the manuscript; gave final approval of the version to be published; and has agreed to be accountable for all aspects of the work.
- AWK: quality checked data; had contributions to the interpretation of the qualitative data; critically commented on versions of the manuscript; gave final approval of the version to be published; and has agreed to be accountable for all aspects of the work.
- DML: had substantial contributions to the design of the work (helped develop protocol and interview questions); reviewed analysis; critically commented on versions of the manuscript; gave final approval of the version to be published; and has agreed to be accountable for all aspects of the work.
- MGP: had substantial contributions to the design of the study (recruited participants); reviewed analysis; critically commented on versions of the manuscript; gave final approval of the version to be published; and has agreed to be accountable for all aspects of the work.
- LN: had substantial contributions to the design of the work (reviewed interview questions); reviewed analysis; critically commented on versions of the manuscript; gave final approval of the version to be published; and has agreed to be accountable for all aspects of the work.
- JG: had substantial contributions to the design of the work (reviewed interview questions); reviewed analysis; critically commented on versions of the manuscript; gave final approval of the version to be published; and has agreed to be accountable for all aspects of the work.
- ICM had substantial contributions to the design of the work (reviewed interview questions); reviewed analysis; critically commented on versions of the manuscript; gave final approval of the version to be published; and has agreed to be accountable for all aspects of the work.
- RS: had substantial contributions to the design of the work (reviewed interview questions); reviewed analysis; critically commented on versions of the manuscript; gave final approval of the version to be published; and has agreed to be accountable for all aspects of the work.
- CB: had substantial contributions to the design of the work (reviewed interview questions); reviewed analysis; critically commented on versions of the manuscript; gave final approval of the version to be published; and has agreed to be accountable for all aspects of the work.
- MP: contributed to the design of the work (reviewed interview questions); reviewed analysis; critically commented on versions of the manuscript; gave final approval of the version to be published; and has agreed to be accountable for all aspects of the work.
- LS: contributed to the design of the work (reviewed interview questions); reviewed analysis; critically commented on versions of the manuscript; gave final approval of the version to be published; and has agreed to be accountable for all aspects of the work.
- EP: contributed to the design of the work (reviewed interview questions); reviewed analysis; critically commented on versions of the manuscript; gave final approval of the version to be published; and has agreed to be accountable for all aspects of the work. CAMM: had substantial contributions to the design of the work (helped develop protocol, data collection schedules); reviewed analysis; contributed to drafting the manuscript; commented on versions of the manuscript; gave final approval of the version to be published; and has agreed to be accountable for all aspects of the work.

## Data Sharing Statement

Data available upon reasonable request.

