## Supplementary 1 for "Attitudes Towards Coronavirus (COVID-19) Vaccine and Sources of Information Across Diverse Ethnic Groups in the UK: a Qualitative Study"

#### Authorship and Affiliations

1. Eirwen Sides (Lead and Corresponding Author)
   a. BSc, Research Assistant
   b. Primary Care and Interventions Unit, Public Health England, Twyver House, Bruton Way, Gloucester, GL1 1PB
   c.
   e. <https://orcid.org/0000-0002-9414-5037>
2. Leah Ffion Jones, PhD, Behavioural Science Team Leader, Behavioural Science and Insights Unit, Public Health England <https://orcid.org/0000-0002-0448-3471>
3. Atiya Kamal, PhD, Senior Lecturer in Health Psychology, School of Social Sciences, Birmingham City University <https://orcid.org/0000-0002-6651-6400>
4. Amy Thomas, MSc, Research Assistant, Primary Care and Interventions Unit, Public Health England <https://orcid.org/0000-0003-2425-0699>
5. Rowshonara B Syeda, MSc, Research Assistant, Primary Care and Interventions Unit, Public Health England
6. Awatif Kaissi, MSc, Research Assistant, Primary Care and Interventions Unit, Public Health England
7. Donna M Lecky, PhD, Head of Unit, Primary Care and Interventions Unit, Public Health England <https://orcid.org/0000-0002-1223-9356>
8. Mahendra G Patel, PhD FRPharmS FHEA, Alumni Fellow NICE, Honorary Visiting Professor, University of Bradford
9. Laura B Nellums, PhD, Assistant Professor in Global Health, Faculty of Medicine & Health Sciences, University of Nottingham, <https://orcid.org/0000-0002-2534-6951>
10. Jane Greenway, Health Practitioner, Public Health England
11. Ines Campos-Matos, Consultant Epidemiologist and Head of Inclusion Health, Public Health England <https://orcid.org/0000-0002-6480-4310>
12. Rashmi Shukla, Director for Midlands and East Region, Public Health England
13. Colin Stewart Brown, Head of Healthcare Associated Infections and Antimicrobial Resistance Division, National Infection Service, Public Health England
14. Manish Pareek, Associate Clinical Professor in Infectious Diseases, Department of Respiratory Sciences, University of Leicester
15. Loretta Sollars, Head of Health Inequalities & Deputy Head Children, Young People and Families, Public Health England <https://orcid.org/0000-0002-4751-029X>
16. Emma Pawson, Head of Regional Engagement and Strategy, Public Health England
17. Cliodna AM McNulty, FRCPathol, Head of Unit to December 2020, (now retired) Primary Care and Interventions Unit, Public Health England <https://orcid.org/0000-0003-4969-5360>

#### Abstract

##### Objectives

To explore attitudes and intentions towards COVID-19 vaccination, and influences and sources of information about COVID-19 across diverse ethnic groups (EGs) in the UK.

##### Design

Remote qualitative interviews and focus groups (FGs) conducted June-October 2020 before UK COVID-19 vaccine approval. Data were transcribed and analysed through inductive thematic analysis.

##### Setting

General public in the community across England and Wales.

##### Participants

100 participants from 19 self-identified EGs with spoken English or Punjabi.

##### Results

Mistrust and doubt were common themes across all EGs including white British and minority EGs, but more pronounced amongst Bangladeshi, Pakistani, Black ethnicities and Travellers. Many participants shared concerns about perceived lack of information about COVID-19 vaccine safety, efficacy and potential unknown adverse effects. Across EGs participants stated occupations with public contact, older adults and vulnerable groups should be prioritised for vaccination. Perceived risk, social influences, occupation, age, co-morbidities and engagement with healthcare influenced participants’ intentions to accept vaccination once available; all Jewish FG participants intended to accept, while all Traveller FG participants indicated they probably would not.

Facilitators to COVID-19 vaccine uptake across all EGs included: desire to return to normality and protect health and wellbeing; perceived higher risk of infection; evidence of vaccine safety and efficacy; vaccine availability and accessibility.

COVID-19 information sources were influenced by social factors, culture and religion and included: friends, family; media and news outlets; and research literature. Participants across most different EGs were concerned about misinformation or had negative attitudes towards the media.

##### Conclusions

During vaccination programme roll-out, including boosters, commissioners and vaccine providers should provide accurate information, authentic community outreach, and use appropriate channels to disseminate information and counter misinformation. Adopting a context-specific approach to vaccine resources, interventions and policies and empowering communities has potential to increase trust in the programme.

### Interview schedule

All participants should have received a copy of the information form and consent form prior to the discussion. Ask all participants to confirm that they have received both and have electronically returned a signed consent form. Ask participants to also confirm on the recording that they are happy to continue with the discussion. Continue to read the paragraph below and allow for people to ask any questions.

#### Introduction [please read this to the interviewee before the interview takes place]

My name is Leah Jones, and I am interviewing you on behalf of Public Health England. We are a government organisation that aim to protect and improve the nation’s health and wellbeing. Knowing your experiences is very important because it means we can offer more support that is helpful in relation to COVID-19, among different groups in society such as different cultures, ages, faiths, religions etc that are living in England.

If you don’t mind, the discussion will be recorded and I will take a few notes. The notes and the recording will be kept completely private, meaning no names or identifiable information that you mention in the recording will be used when it is typed up, meaning we will not use your name or any other information that could be used to identify you, having said that, you may want to stick to first names and avoid using identifiable information for the sake of the recording, but this is entirely your choice. Are you happy to go ahead with the discussion?

#### Background Questions

Can I ask that we go around the group and introduce ourselves? It would be useful to know your name and whether you are first, second, third, fourth generation.

#### COVID-19 questions, general

1. Tell me about your experiences of COVID-19. *(Environmental context and resources) Probe: Have you or someone you know had it,*
2. How has the COVID-19 pandemic made you feel? *(Emotion)*
3. How do you think your experience is different because you come from a BAME/minority ethnic background? *(Professional role and identity)*
4. Tell me about how the pandemic may have impacted on the support structures within your circles of friends and family. *(Social norms, Environmental context and resources) Probe: Have you been able to give/receive support in the way you normally would? How has it changed if at all?*
5. Is there anything you will do differently as a result of this pandemic? *Probe: hand washing, hygiene*, *self-care, GP visits, antibiotics, left-overs, medication, (Intentions)*

#### Self-care

1. Tell me about how you would self-care/look after yourself at home if you developed COVID-19 symptoms. *(Skills)*

#### Health seeking

2. When would you seek a consultation with a healthcare professional if you suspect yourself of having COVID-19? *(Memory, attention and decision making)*
3. How important is it to you to consult with a healthcare professional if you suspect COVID-19 in yourself or a family member? *(Environmental context and resources)*
4. What would you expect to happen in a consultation for suspected COVID-19? *(Goals, beliefs about consequences)* *Probe: antibiotics, other medication, reassurance, advice etc.*

#### Prevention

5. Tell me about any strategies you have used to try and prevent yourself from catching COVID-19. *Probe: hand washing, face coverings, not touching face, avoiding social situations* *(Skills)*
6. How confident are you that those strategies will/have worked? *(Beliefs about capabilities)*

#### Vaccines

7. How optimistic are you that a vaccine will help solve the pandemic? *(optimism)*
8. How would you feel about being offered a vaccine for COVID-19 if and when one becomes available? *(Emotion) (Memory, attention and decision making)*
9. Tell me why you would or would not accept a COVID-19 vaccine. *(Intentions)*
   a. Probe: Would cost influence your decision? *(Memory, attention and decision making)*
   b. Would effectiveness influence your decision? e.g. if it only lasted a year or so? *(Memory, attention and decision making)*
10. Who do you think should receive the COVID-19 vaccine, and why? Probe: children, elderly, health workers, essential workers, all *(Professional role and identity)*
11. If you were willing to receive a vaccine, where would you be happy to receive the COVID-19 vaccine? E.g. Pharmacy, GP, hospital, schools, specialist vaccine sites. *(Environmental context and resources)*
    a. Is there anywhere you wouldn’t visit to receive a vaccine?

#### Testing

12. Have you received a test for COVID-19?
13. What are your thoughts on receiving an antibody test (Test to see if you’ve already had it)? *(Memory, attention and decision making)*
14. When would you like to receive a test? At the start of symptoms? After the illness? *(Environmental context and resources)*
15. Who do you think should be tested? Probe: children, elderly, health workers, essential workers, all *(Professional role and identity)*
16. Where would you be happy to receive a COVID-19 test? E.g. Pharmacy, GP, hospital, schools, specialist testing sites. *(Environmental context and resources)*
    b. Is there anywhere you wouldn’t visit to receive a test?

#### Shielding/at risk groups

17. To what extent would you consider yourself at risk? *(Professional role and identity, beliefs about consequences)*
18. What do you think of the recent figures showing higher mortality rates among BAME groups? *(Beliefs about consequences) (Environmental context and resources)*
    a. To what extent has this affected your behaviour? *(Memory, attention and decision making)*
19. Tell me about any experiences you have of family members at increased risk or are shielding. *(Environmental context and resources)*
20. How has this made you feel? *(Emotion)*
21. Have you done anything differently for those individuals? If so, what? *(Environmental context and resources)*
22. How easy or difficult have you found shielding? *(Beliefs about capabilities)*
23. What do you think will happen to those vulnerable groups now that lockdown is being eased? *(Beliefs about consequences)*

#### Government messaging

24. What is your experience of the government health messages telling people what to do during this pandemics?
    c. Is there anything that you’ve found difficult about understanding what to do during this pandemic? *(Memory, attention and decision making) probe: about specific messages: stay at home; keep safe; save the NHS. Stay alert; control the virus; save lives.*
    d. Is there anything that you’ve found easy about understanding what to do during this pandemic? *(Memory, attention and decision making) probe: about specific messages: stay at home; keep safe; save the NHS. Stay alert; control the virus; save lives.*
25. If you need information about COVID-19, where would you go to get it? *Probe: Website? Family? Pharmacy?* *(Environmental context and resources)*

Interviewer to go through the different messages and discuss each one.

26. How easy or difficult have you found physical distancing? *(Beliefs about capabilities)*
27. How will you decide when to start seeing your friends and family again? Probe: When the government says, practical reasons, when you want etc. *(Memory, attention and decision making)*
28. Have you broken any of the guidelines issued by the government, if so, what did you do and why? (Re-assure participants of confidentiality and anonymity to ensure honest answers) *(Environmental context and resources)*

#### Returning to work/schools

29. What are your thoughts on children going back to schools?
    c. Emotions: fear, worried, relieved, happy *(Emotion)*
    d. Concerns: travel, anxiety, distancing in schools, hygiene *(Environmental context and resources)*
30. What are your thoughts on returning back to work, if applicable?
    e. Emotions: fear, worried, relieved, happy *(Emotion)*
    f. Concerns: travel, risk, family, caring *(Environmental context and resources)*
31. Based on our discussions, what do you feel you need, moving forwards? Probe: guidance, testing, vaccine, medication, *(Environmental context and resources)*
