## Supplementary 2 for "Attitudes Towards Coronavirus (COVID-19) Vaccine and Sources of Information Across Diverse Ethnic Groups in the UK: a Qualitative Study"

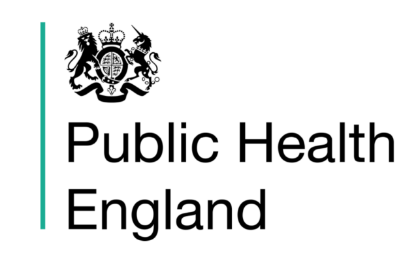

### Take part in a 60 minute group discussion and earn £25

Public Health England are looking for individuals to take part in **60 minute** discussions all around experiences of the **COVID-19 pandemic**.

We are specifically looking to recruit individuals from **minority ethnic backgrounds**.

Discussions will focus on your experiences of COVID-19, such as physical distancing, your perceptions of vaccines and testing, shielding at risk individuals, how you’ve coped and managed under the government guidelines and your health related behaviours.

The aim of this work is to inform interventions to help minority ethnic groups during this pandemic, and to also inform the development of a general self-care leaflet.

**We are offering everyone who takes part £25 for their time.**

If you are interested in taking part, or would like more information, please contact the lead researcher at the following details:

**Leah Jones, Research Project Support Officer, Public Health England**

**+442084953256**

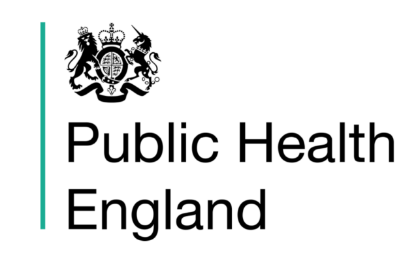

### Take part in a 60 minute group discussion and earn £25

Public Health England are looking for individuals to take part in **60 minute** discussions all around experiences of the **COVID-19 pandemic**. We are interested in understanding how experiences may be different across different ethnic groups.

For the next focus group we are specifically looking to recruit **white British individuals** from a range of ages, genders and backgrounds.

Discussions will focus on your experiences of COVID-19, such as physical distancing, how you’ve coped and managed under the government guidelines and your health-related behaviours.

**We are offering everyone who takes part £25 for their time.**

If you are interested in taking part, or would like more information, please contact the lead researcher at the following details:

**Leah Jones, Research Project Support Officer, Public Health England**

**+442084953256**
