## Supplementary 3 for "Attitudes Towards Coronavirus (COVID-19) Vaccine and Sources of Information Across Diverse Ethnic Groups in the UK: a Qualitative Study"

**Supplementary 4.** Focus group ethnicity and gender breakdown

| **Focus group (FG) name and gender of participants** | **Self-reported ethnicity breakdown** | **n** |
| --- | --- | --- |
| FG1  5F, 2M | Bangladeshi | 2 |
|  | Indian/Mauritian | 1 |
|  | Latin American | 1 |
|  | Pakistani | 1 |
|  | Sri Lankan | 1 |
|  | Unknown | 1 |
| FG2  4F | Asian | 1 |
|  | Black African | 1 |
|  | Latin American | 1 |
|  | Pakistani | 1 |
| FG3  1F, 3M | Bangladeshi | 2 |
|  | Black Caribbean | 1 |
|  | Chinese | 1 |
| FG4  3F, 1M | Bangladeshi | 3 |
|  | Sri Lankan | 1 |
| FG5  2F, 1M | Asian | 1 |
|  | Black African | 1 |
|  | Pakistani | 1 |
| FG6  4F, 1M | Pakistani | 5 |
| FG7  1F, 1M, 1 Unknown | Chinese | 3 |
| FG8  2F, 3M | Black African | 3 |
|  | Black Caribbean | 2 |
| FG9  2F, 4M | Indian | 5 |
|  | Indian/Mauritian | 1 |
| FG10  3F, 2M | Bangladeshi | 5 |
| FG11  1F, 1M | Lithuanian | 1 |
|  | Polish | 1 |
| FG12  2F, 2M | Chinese | 4 |
| FG13  2F | Arabic | 2 |
| FG14  1F, 5M | Indian | 5 |
|  | Vietnamese | 1 |
| FG15  3M | Bangladeshi | 3 |
| FG16  2F, 2M | Black | 2 |
|  | Black African | 1 |
|  | Black British | 1 |
| FG17  1F, 1M | Chinese | 2 |
| FG18  1F, 3M | Indian | 4 |
| FG19  3M | Bangladeshi | 1 |
|  | Pakistani | 2 |
| FG20  1F, 3M | White British | 4 |
| FG21  3F, 4M | Jewish | 7 |
| FG22  1F, 2M | White British | 3 |
| FG23  1F, 2M | Black African | 3 |
| FG24  3F, 1M | Traveller | 4 |
| Interview 1  1F | Pakistani | 1 |
| Interview 2  1F | Pakistani | 1 |
| Interview 3  1F | Pakistani | 1 |
