## Supplementary 4 for "Attitudes Towards Coronavirus (COVID-19) Vaccine and Sources of Information Across Diverse Ethnic Groups in the UK: a Qualitative Study"

SRQR Reporting checklist for qualitative study.

**Instructions to authors**

Complete this checklist by entering the page numbers from your manuscript where readers will find each of the items listed below. SRQR reporting guidelines: O'Brien BC, Harris IB, Beckman TJ, Reed DA, Cook DA. Standards for reporting qualitative research: a synthesis of recommendations. Acad Med. 2014;89(9):1245-1251.

| No. | Topic | Reporting Item | Page no. |
| --- | --- | --- | --- |
|  | **Title and abstract** |  |  |
| S1 | Title | Concise description of the nature and topic of the study Identifying  the study as qualitative or indicating the approach (e.g., ethnography,  grounded theory) or data collection methods (e.g., interview, focus  group) is recommended | 1 |
| S2 | Abstract | Summary of key elements of the study using the abstract format of  the intended publication; typically includes background, purpose,  methods, results, and conclusions | 1-2 |
|  | **Introduction** |  |  |
| S3 | Problem formulation | Description and significance of the problem/phenomenon studied;  review of relevant theory and empirical work; problem statement | 3 |
| S4 | Purpose of research or question | Purpose of the study and specific objectives or questions | 3 |
|  | **Methods** |  |  |
| S5 | Qualitative approach and research paradigm | Qualitative approach (e.g., ethnography, grounded theory, case study,  phenomenology, narrative research) and guiding theory if appropriate;  identifying the research paradigm (e.g., postpositivist, constructivist/  interpretivist) is also recommended; rationale | 3 |
| S6 | Research characteristics and reflexivity | Researchers’ characteristics that may influence the research, including  personal attributes, qualifications/experience, relationship with  participants, assumptions, and/or presuppositions; potential or actual  interaction between researchers’ characteristics and the research  questions, approach, methods, results, and/or transferability | 4 |
| S7 | Context | Setting/site and salient contextual factors; rationale | 3-4 |
| S8 | Sampling strategy | How and why research participants, documents, or events were  selected; criteria for deciding when no further sampling was necessary  (e.g., sampling saturation); rationale | 3-4 |
| S9 | Ethical issues pertaining to human subjects | Documentation of approval by an appropriate ethics review board  and participant consent, or explanation for lack thereof; other  confidentiality and data security issues | 4 |
| S10 | Data collection methods | Types of data collected; details of data collection procedures including  (as appropriate) start and stop dates of data collection and analysis,  iterative process, triangulation of sources/methods, and modification  of procedures in response to evolving study findings; rationale | 4 |
| S11 | Data collection instruments and technologies | Description of instruments (e.g., interview guides, questionnaires)  and devices (e.g., audio recorders) used for data collection; if/how the  instrument(s) changed over the course of the study | 4 |
| S12 | Units of study | Number and relevant characteristics of participants, documents, or  events included in the study; level of participation (could be reported  in results) | 5 |
| S13 | Data processing | Methods for processing data prior to and during analysis, including  transcription, data entry, data management and security, verification  of data integrity, data coding, and anonymization/deidentification of  excerpts | 4 |
| S14 | Data analysis | Process by which inferences, themes, etc., were identified and  developed, including the researchers involved in data analysis; usually  references a specific paradigm or approach; rationale | 4 |
| S15 | Techniques to enhance trustworthiness | Techniques to enhance trustworthiness and credibility of data analysis  (e.g., member checking, audit trail, triangulation); rationale | 3-4 |
|  | **Results/Findings** |  |  |
| S16 | Synthesis and interpretation | Main findings (e.g., interpretations, inferences, and themes); might  include development of a theory or model, or integration with prior  research or theory | 5-18 |
| S17 | Links to empirical data | Evidence (e.g., quotes, field notes, text excerpts, photographs) to  substantiate analytic findings | 5-18 and supplementary files |
|  | **Discussion** |  |  |
| S18 | Integration with prior work, implications,  transferability, and contribution(s) to the field | Short summary of main findings; explanation of how findings  and conclusions connect to, support, elaborate on, or challenge  conclusions of earlier scholarship; discussion of scope of application/  generalizability; identification of unique contribution(s) to scholarship  in a discipline or field | 19-24 |
| S19 | Limitations | Trustworthiness and limitations of findings | 20-21 |
|  | **Other** |  |  |
| S20 | Conflicts of interest | Potential sources of influence or perceived influence on study conduct  and conclusions; how these were managed | 25 |
| S21 | Funding | Sources of funding and other support; role of funders in data  collection, interpretation, and reporting | 25 |

The SRQR checklist is distributed with permission of Wolters Kluwer © 2014 by the Association of American Medical Colleges.

This checklist can be completed online using https://www.goodreports.org/, a tool made by the EQUATOR Network in collaboration with Penelope.ai
